# A multiplexed high-throughput neutralization assay reveals a lack of activity against multiple variants after SARS-CoV-2 infection

**DOI:** 10.1101/2021.04.08.21255150

**Authors:** Craig Fenwick, Priscilla Turelli, Céline Pellaton, Alex Farina, Jérémy Campos, Charlène Raclot, Florence Pojer, Valeria Cagno, Giuseppe Pantaleo, Didier Trono

## Abstract

The detection of SARS-CoV-2-specific antibodies in the serum of an individual indicates prior infection or vaccination. However, it provides limited insight into the protective nature of this immune response. Neutralizing antibodies recognizing the viral Spike are far more revealing, yet their measurement traditionally requires virus- and cell-based systems that are costly, time-consuming, poorly flexible and potentially biohazardous. Here we present a cell-free quantitative neutralization assay based on the competitive inhibition of trimeric SARS-CoV-2 Spike protein binding to the angiotensin converting enzyme 2 (ACE2) viral receptor. This high-throughput method matches the performance of the gold standard live virus infectious assay, as verified with a panel of 206 seropositive donors with varying degrees of infection severity and virus-specific IgG titers, achieving 96.7% sensitivity and 100% specificity. Furthermore, it allows for the parallel assessment of neutralizing activities against multiple SARS-CoV-2 Spike variants of concern (VOC), which is otherwise unpredictable even in individuals displaying robust neutralizing antibody responses. Profiling serum samples from 59 hospitalized COVID-19 patients, we found that although most had high activity against the 2019-nCoV Spike and to a lesser extent the B.1.1.7 variant, only 58% could efficiently neutralize a Spike derivative containing mutations present in the B.1.351 variant. In conclusion, we have developed an assay that has proven its clinical relevance in the large-scale evaluation of effective neutralizing antibody responses to VOC after natural infection and that can be applied to the characterization of vaccine-induced antibody responses and of the potency of human monoclonal antibodies.

**Once sentence summary:** Multiplexed cell-free neutralization assay for quantitative assessment of serum antibody responses against Spike mutations in SARS-COV-2 variants

## Main

The severe acute respiratory syndrome coronavirus 2 (SARS-CoV-2) pandemic is causing a global crisis with a devastating impact on public health, societies and economies around the world.*(1, 2)* The majority of individuals infected with SARS-CoV-2 experience mild to moderate symptoms that do not require hospitalization but risk factors including age, ethnicity, gender, obesity and underlying health issues including cardiovascular disease, diabetes and chronic respiratory disease can lead to severe illness. *(3)* COVID-19, the disease caused by SARS-CoV-2 infection, has so far resulted in >2.5 million deaths worldwide and it is estimated that approximately 5-10% of symptomatic infected individuals will have longer term health consequences.*(4)*

The global spread of SARS-CoV-2 led to the rapid development of diagnostic tools including viral detection tests and serological assays to assist in the public health management of the pandemic.*(5, 6)* Seroprevalence studies generally search for the presence of virus-specific antibodies in the serum of individuals as a marker of previous infection or, albeit still too rarely, vaccination. However, these analyses do not assess whether the detected immune response is protective.*(7)* Indeed, only a subset of antibodies mounted against a virus can block its spread. These are called neutralizing antibodies, and in the case of SARS-CoV-2 they recognize Spike (S), the viral surface protein responsible for mediating entry through binding of the ACE2 viral receptor.*(8) (9)*

The gold standard for measuring levels of SARS-CoV-2-specific neutralizing antibodies relies on infection of ACE2-expressing cells with live virus, monitoring the reduction in the culture of virus-induced cytopathic effects (CPE) or, if a virus modified to encode GFP or luciferase is used, the expression of these reporters. *(10)* Such neutralization assays are not routinely performed because they are technically demanding, take several days for readout and require trained professionals working in biosafety level 3 (BSL3) facilities. As an alternative, viral pseudotypes can be utilized, typically lentiviral vector (LV) or vesicular stomatitis virus (VSV) particles coated with the SARS-CoV-2 S protein to drive their entry into ACE2-positive target cells. Although compatible with the less constrained environment of biosafety level 2 laboratories, these assays still require cell culture and take several days.*(11)* Furthermore, neither LV nor VSV pseudotypes fully recapitulate SARS-CoV-2 infectivity, because their virions assemble at the plasma membrane whereas the coronavirus loads its S protein in the endoplasmic reticulum-Golgi intermediate compartment.*(12)* As a result, these S-pseudotyped virions are difficult to produce as highly infectious particles and are inherently easier to neutralize compared to the authentic SARS-CoV-2 virus, leading to often erratic estimations of the neutralizing activity of biological samples.

Measuring the neutralizing activity of virus-specific antibodies is further complicated by the continuous emergence of SARS-CoV-2 variants. Some of these variants are of particular clinical relevance, such as ones with mutations in and around the receptor binding domain (RBD) of the S protein, resulting in increased affinity for the ACE2 receptor or reduced recognition by neutralizing antibodies.*(13, 14)* For instance, the B.1.351 (also called 501Y.V2 or South African variant of concern (VOC)) has been found largely to escape immunity induced by some COVID-19 vaccines,*(15)* and the closely related P.1 (also called 501Y.V3, B.1.1.28 or Brazilian VOC) to be responsible for large numbers of reinfections in the Manaus region. *(16)*

To address these challenges, we developed a cell-free neutralization assay based on the competitive inhibition of ACE2 binding to S protein trimers-bearing beads. This method is high-throughput, quantitative, yields results tightly correlating those obtained with a classical wild-type virus cell-based neutralization assay, and allows the simultaneous evaluation of multiple S protein variants. Illustrating its value, it reveals that some previously infected individuals display variant-specific serum neutralizing antibody titers, suggesting that they remain at risk of infection with at least some VOCs. Based on these results, we propose that this novel assay stands to play an important role in analyzing the protection induced by SARS-CoV-2 infection or vaccination, thus facilitating the determination and certification of individuals’ antiviral immunity status and the adaptation of vaccines to the potential emergence of escape variants.

## Results

### A cell-free SARS-CoV-2 S protein trimer /ACE2-based neutralization assay

SARS-CoV-2 uses angiotensin-converting enzyme 2 (ACE2) as its primary receptor, which it recognizes via the receptor binding domain (RBD) of its S protein.*(17)* In return, this 211 amino acid-long region is the main target of neutralizing antibodies.*(8, 18, 19)* We thus hypothesized that an assay capable of providing a quantitative measurement of the inhibition of SARS-CoV-2 S protein interaction with ACE2 could reveal the neutralizing potential of antibodies found in serum and other biological samples. However, whereas a previously described system based on this assumption relied on the sole use of S protein RBD monomers,*(20)* we reasoned that having the full S protein in trimeric higher order structure, as found on the surface of the virion, was more likely to recapitulate the physiological configuration. *(21)* Therefore, we produced S protein trimers (S^3^) in native prefusion configuration *(22)* in CHO cells and coupled these proteins to Luminex beads. We then measured the ability of these S^3^-bearing beads to recruit a recombinant human ACE2 mouse Fc-tagged fusion protein (ACE2-Fc), which we detected with a fluorescently tagged anti-mouse Fc secondary antibody. A standard protocol was then established (**Figure 1a**), whereby after S^3^-carrying beads are first mixed in a 96-well plate with limited dilutions of test serums, before adding ACE2-Fc and pursuing incubation for 60 min in order to reach equilibrium, and finally measuring the amount of captured soluble viral receptor with a fluorescently tagged antibody on a Bio-Plex 200 System. Using this procedure, we first verified that serum samples from pre-COVID-19 pandemic healthy donors (n=104, **Figure 1b-c**) did not significantly cross-react *(2)* and interfere with ACE2-Fc binding to S^3^-coupled beads, whereas serum from post-infected donors induced a dilution-dependent signal reduction (**Figure 1d**).

**Figure 1:**
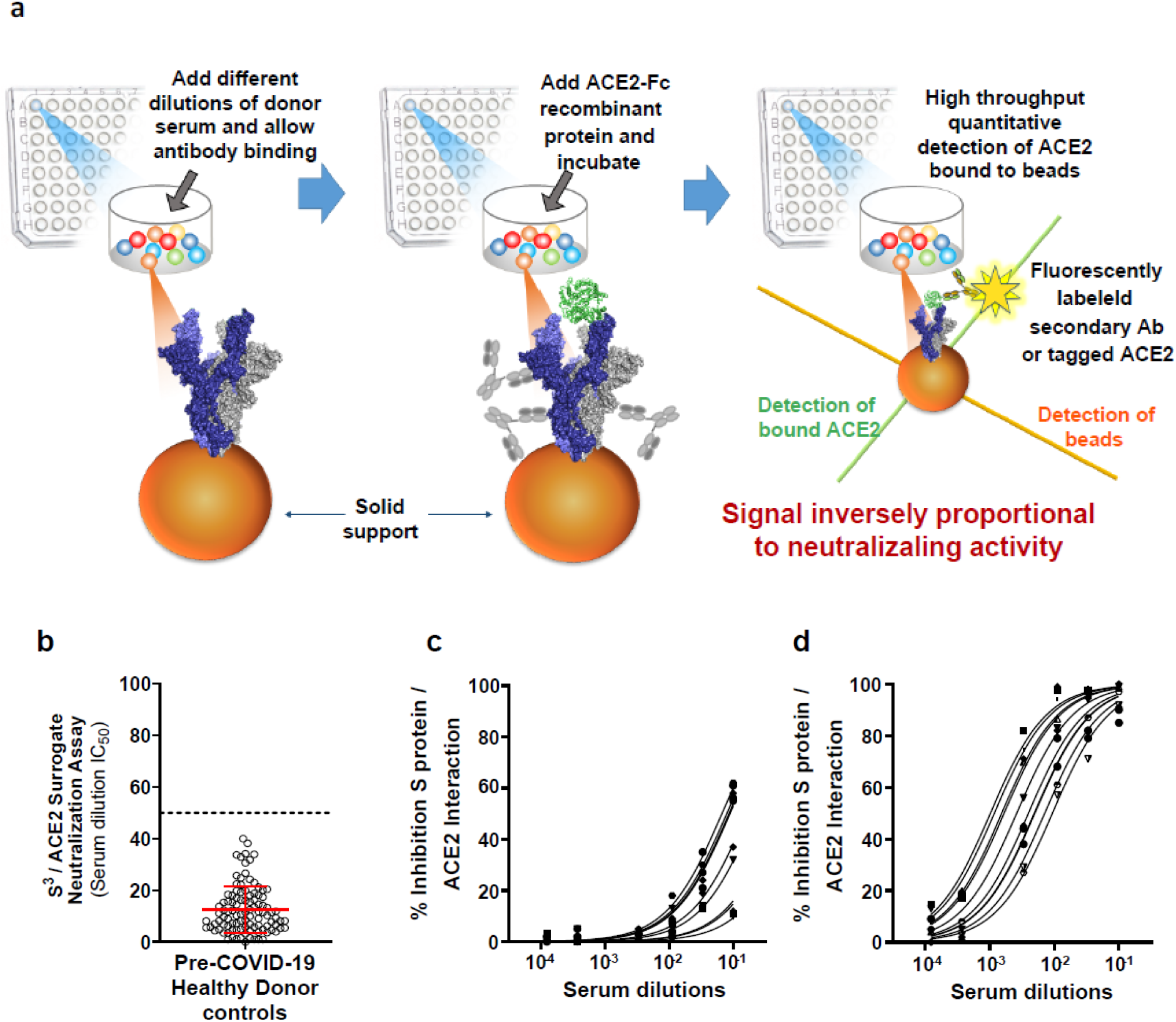
Outline and validation of a cell-free SARS-CoV-2 S protein trimer /ACE2 surrogate neutralization assay. (**a**) Schematic outline of the S^3^ /ACE2 neutralization assay. Anti-SARS-CoV-2 serum antibodies are monitored for their capacity in blocking the S^3^ /ACE2 interaction. ACE2 binding to S protein was detected though use of a fluorescently labeled secondary antibody and the signal intensities are inversely proportional to the neutralizing potential of the anti-S protein antibodies. (**b**) Serum dilution IC_50_ values were calculated for 104 healthy adult donor samples collected prior to November 2019 (pre-COVID-19 pandemic). The mean IC50 values and SD were used to establish a lower limit cutoff of 50 indicated by the dashed line (mean IC_50_ of 12.5 + 4 × 9.0 SD = 50 serum dilution). (**c**) Representative concentration response curves for ten healthy donor serum samples and (**d**) ten SARS-CoV-2 seropositive donors with varying levels of anti-S protein IgG antibody. Mean ± s.d. shown in graph **b**. S protein / ACE2 structure was generated with PDB 7a98.

We went on to validate the surrogate neutralization assay with a panel of 206 post-infection serums obtained from individuals, some with a prior history of RT-PCR-documented symptomatic infection requiring or not hospitalization (n=95), and others without known SARS-CoV-2 antecedent but identified as seropositive in a Swiss population serological survey (n=111). As expected, an inverse correlation was noted when comparing these two groups between average serum anti-S protein IgG and/or IgA levels and severity of previous disease (**Figure 2a-b**). These 206 serums were tested in parallel with the cell-free S^3^-ACE2 assay and a conventional live CPE assay in Vero cells. The results revealed a high correlation of the neutralizing titers obtained with the two assays (R^2^ = 0.825) over the >3 log range measured amongst the various samples (**Figure 3a**). We additionally subjected a subset of these serums (n=75) to a LV reporter pseudotype-based neutralization assay, which revealed a weaker correlation with the live virus assay (R^2^ = 0.65, n=75; **Figure 3b**).

**Figure 2:**
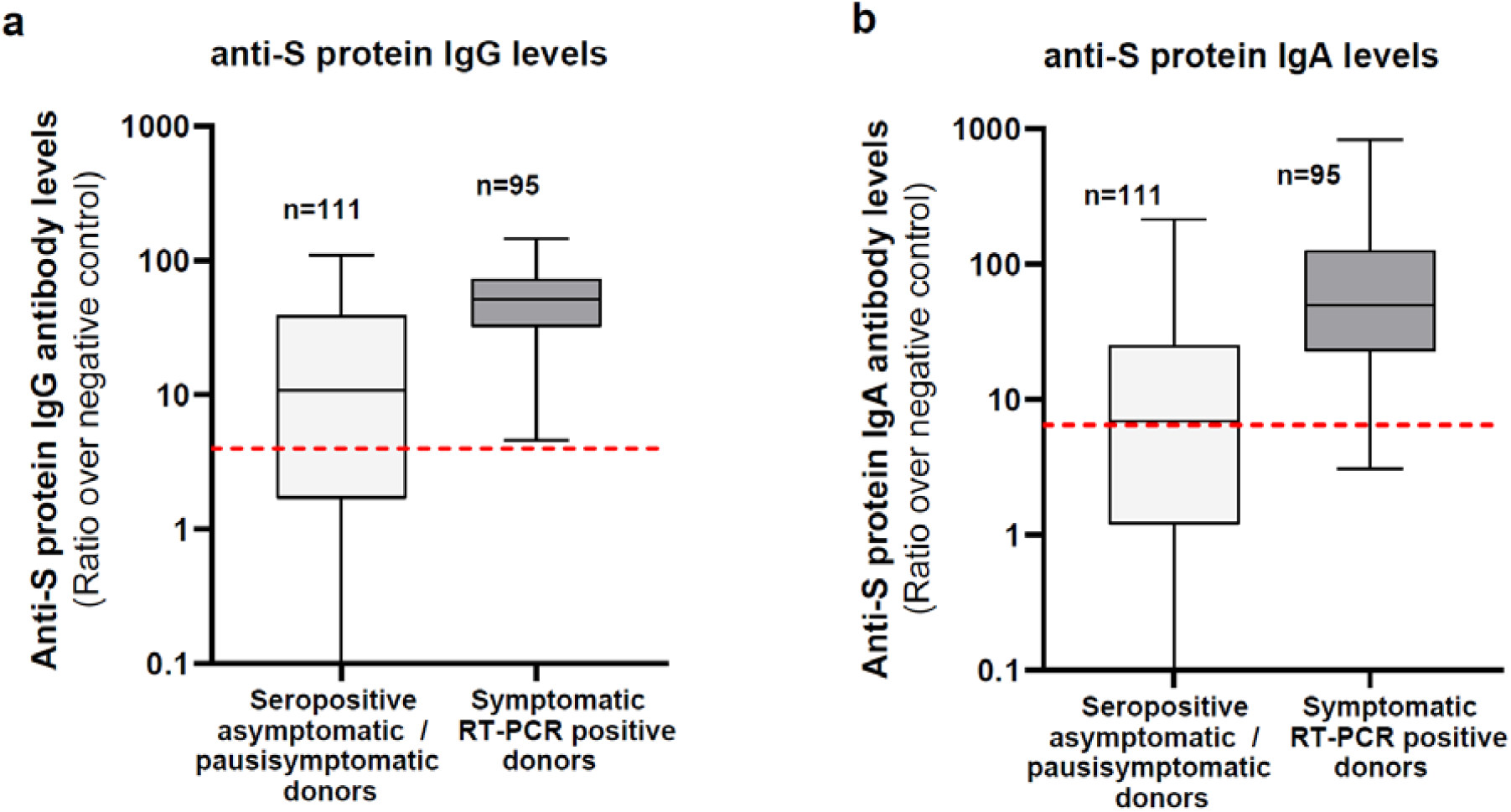
Anti-S protein IgG and IgA levels in the seropositive donor groups. Anti-S protein IgG (**a**) and IgA (**b**) antibody levels measured in the serum samples of different donor groups using a Luminex Spike trimer serological assay. Donors were seropositive for anti-S protein IgG and/or IgA antibodies with positivity cutoffs of 4 and 6.5 ratios over a standard negative control, respectively (red dashed line in (**a**) and (**b**)*(21)*. Donor groups consist of seropositive volunteers, randomly selected individuals from the general population and donors that were in contact with a RT-PCR confirmed infected subject (n=111) with reduced level of symptoms and symptomatic RT-PCR positive donors that were either hospitalized COVID-19 patients or not (n=95). Boxplots shown mean and the 95% interquartile range.

**Figure 3:**
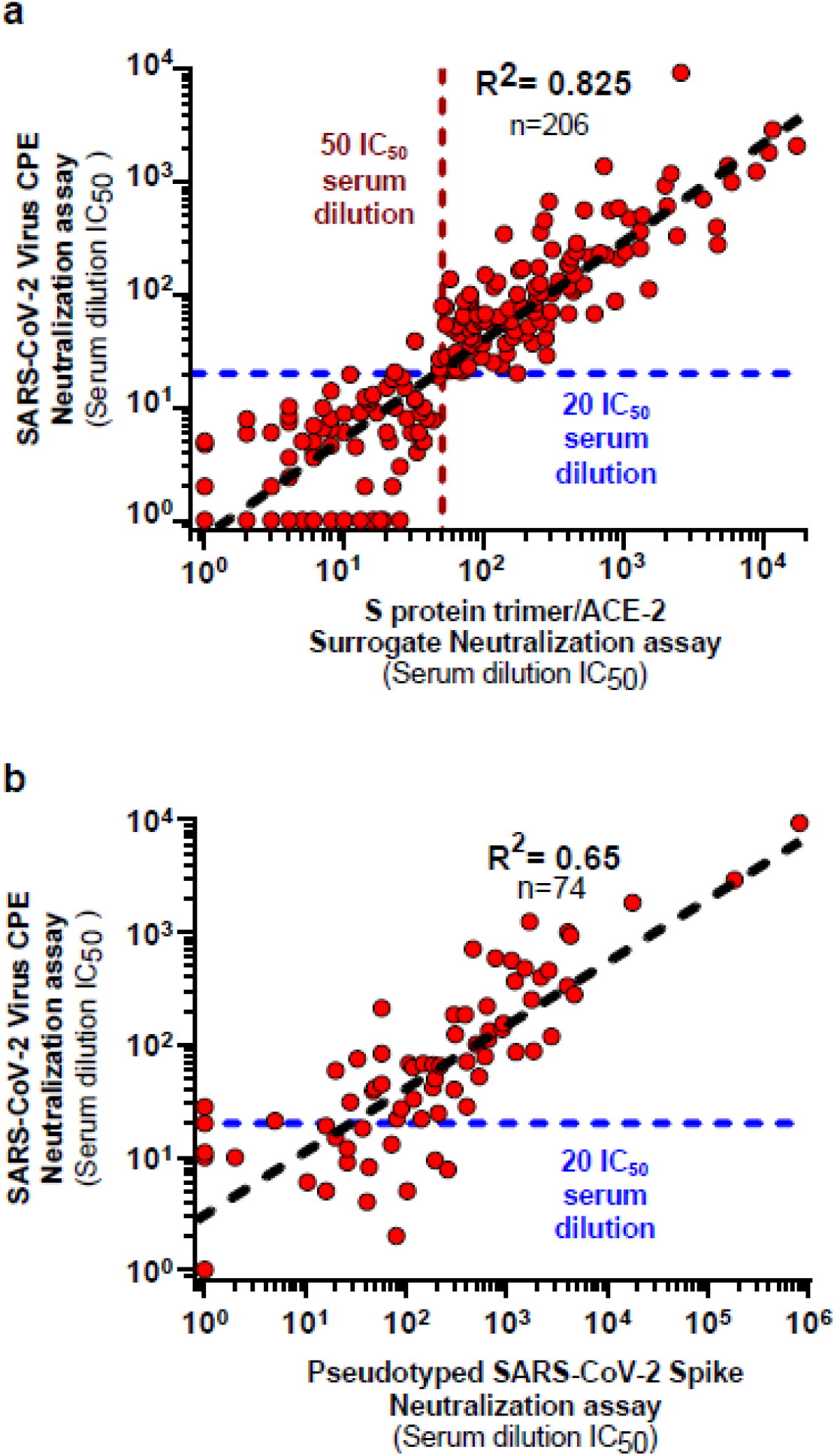
Cross-validation of a cell-free SARS-CoV-2 S protein trimer /ACE2 surrogate neutralization assay. (**a**) Cross-validation studies between S^3^/ACE2 surrogate neutralization assay and the “gold standard” SARS-CoV-2 cytopathic effect (CPE) neutralization assay. Seropositive donors (n=206) with varying levels of anti-S protein antibodies were selected for comparison of the assays. Donors consisted of COVID-19 hospitalized patients (n=31), symptomatic infected donors with RT-PCR-documentation (n=64) and other seropositive donors identified through random sampling, volunteers or contact with a confirmed SARS-CoV-2 infected individual in a Swiss population serological survey (n=111). (**b**) Correlation between the live SARS-CoV-2 virus cytopathic effect and S protein pseudotyped neutralization assays. A group of 74 samples from S protein seropositive donors were compared in the live virus CPE and S protein pseudotyped virus cell based neutralization assays. Correlation between the two neutralization assays is represented by the black dashed line and the 20 serum dilution IC_50_ cutoff for positivity in the CPE assay is shown with the blue dashed line.

For the S^3^-ACE2 neutralization assay, a lower limit IC_50_ serum dilution of 50 was set as the specificity cutoff using IC50 values for the 104 pre-COVID-19 pandemic healthy donor samples (mean IC_50_ of 12.5 + 4 × 9.0 SD; **Figure 1b**). A serum dilution IC_50_ of 20 was selected as the cutoff for positivity in the live SARS-CoV-2 virus CPE assay given that this corresponds to 95-99% viral neutralization (using Nonlinear regression with 1 and 1.5 Hill slopes values to achieve IC_95_ and IC_99_, respectively; **Supplemental Figure 1**) for the undiluted serum sample.*(23)* Using these criteria, the surrogate neutralization assay was determined to achieve 96.7% sensitivity (118 out of 122) and 100% specificity (0 out of 84) relative to the live virus CPE assay (**Table 1**). Additionally, samples with moderate blocking activity in the S^3^-ACE2 neutralization assay (serum dilution 1:50 to 1:100) strongly correlated with samples that exhibited weak (1:20 to 1:50) to moderate (1:50 to 1:100) neutralizing activities in the live virus assay.

**Table 1:** Benchmarked performance of the cell-free S^3^ /ACE2 surrogate neutralization assay against the SARS-CoV-2 live virus CPE neutralization assay.

|  |  | Neutralizing activity in the live virus CPE assay<br>(IC<sub>50</sub> serum dilution) |  |
| --- | --- | --- | --- |
| | | Below lower limit cutoff for<br>neutralizing activity<br>($<20$ IC<sub>50</sub>; n= 84) | Positive neutralizing activity<br>($>20$ IC<sub>50</sub>; n= 122) |
| **S<sup>3</sup> / ACE2**<br>**assay**<br>**(IC<sub>50</sub> serum**<br>**dilution)** | **Below lower limit**<br>**ACE2 blocking cutoff**<br>**($< 50$ IC<sub>50</sub>; n=88)** | n=84 S<sup>3</sup> / n=84 CPE | **3.2% False negative**<br>n=88 S<sup>3</sup> / n=4 CPE |
|  | **Positive for ACE2**<br>**blocking activity**<br>**($>50$ IC<sub>50</sub>; n=118)** | **0% False positive**<br>n=0 S<sup>3</sup> / n=84 CPE | **96.7% Sensitivity**<br>n=118 S<sup>3</sup> / n=122 CPE |

### Multiplexed measurement of SARS-CoV-2 VOCs neutralization

A characteristic of Luminex bead-based assays is the use of analytical optics that allow for the simultaneous evaluation of analytes captured by multiple baits, each coupled to beads of different colors. We took advantage of this feature to test in parallel the neutralization potential of post-infection serums against a large array of SARS-CoV-2 S protein variants. For this, we produced a set of S protein derivatives containing specific mutations alone or in combination, amongst which amino acid changes suspected to contribute to immune escape of some VOCs, such as substitutions at positions K417, E484 and N501 found in the B.1.1.7 (501Y.V1 or UK variant), B.1.351 (501Y.V2 or South African variant) and P.1 strains (501Y.V3 or Brazilian variant) (**Table 2**)*(15, 16, 24)*. We then coupled each of the corresponding S protein trimers to beads of a given color, and placed equal amounts of each of these beads in the wells of a 96-sell plate. To validate the utility of the S^3^-ACE2 assay in identifying S protein mutations conferring reduced sensitivity to neutralization, concentration response curves were generated with the REGN10933 therapeutic antibody profiled against the K417N/E484K/N501Y and Δ69-70 Y453F S protein mutations present in the B.1.351 and mink SARS-CoV-2 variants, respectively (**Figure 4a**). Consistent with published results, REGN10933 had a significant loss in potency against S proteins harboring mutations from either the B.1.351 (61-fold loss in IC_80_ activity) or mink variants (67-fold loss in IC_80_ activity) *(16, 25)*. We then performed the neutralization assay on a large series of post-infection serums, selected examples of which are illustrated in **Figure 4b-c**. In an S protein panel consisting of single amino acid mutations, two donors both exhibited high neutralizing antibody titers (>1,000 IC_50_) against the original 2019-CoV S protein, as measured in duplicates with beads of two different colors. However, while one (sample 3506) retained full potency against a series of seven S protein point mutants, the other (sample 9504) displayed a >2 log reduced ability to block the interaction of ACE2 with the E484K S protein mutant, a result corroborated in the LV pseudotype reporter assay (**Supplemental Figure 2**).

**Table 2:**
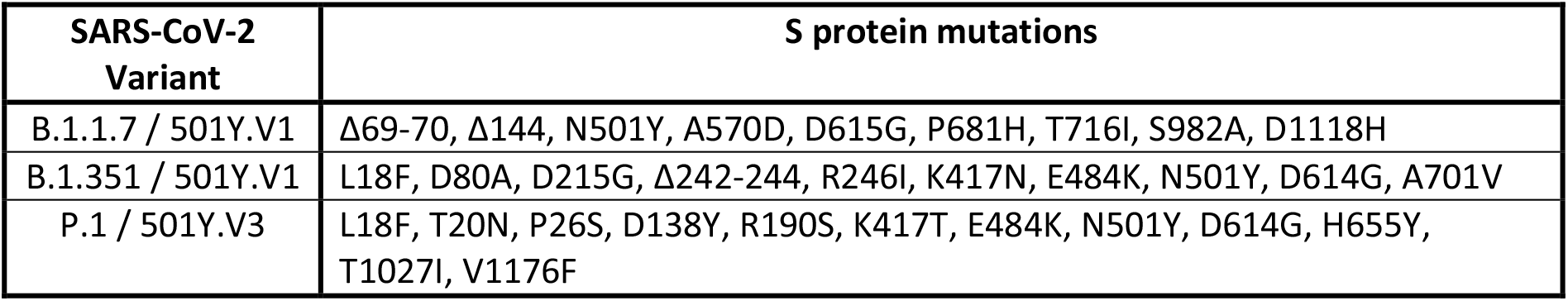
S protein mutations in different circulating SARS-CoV-2 variants of concern

**Figure 4:**
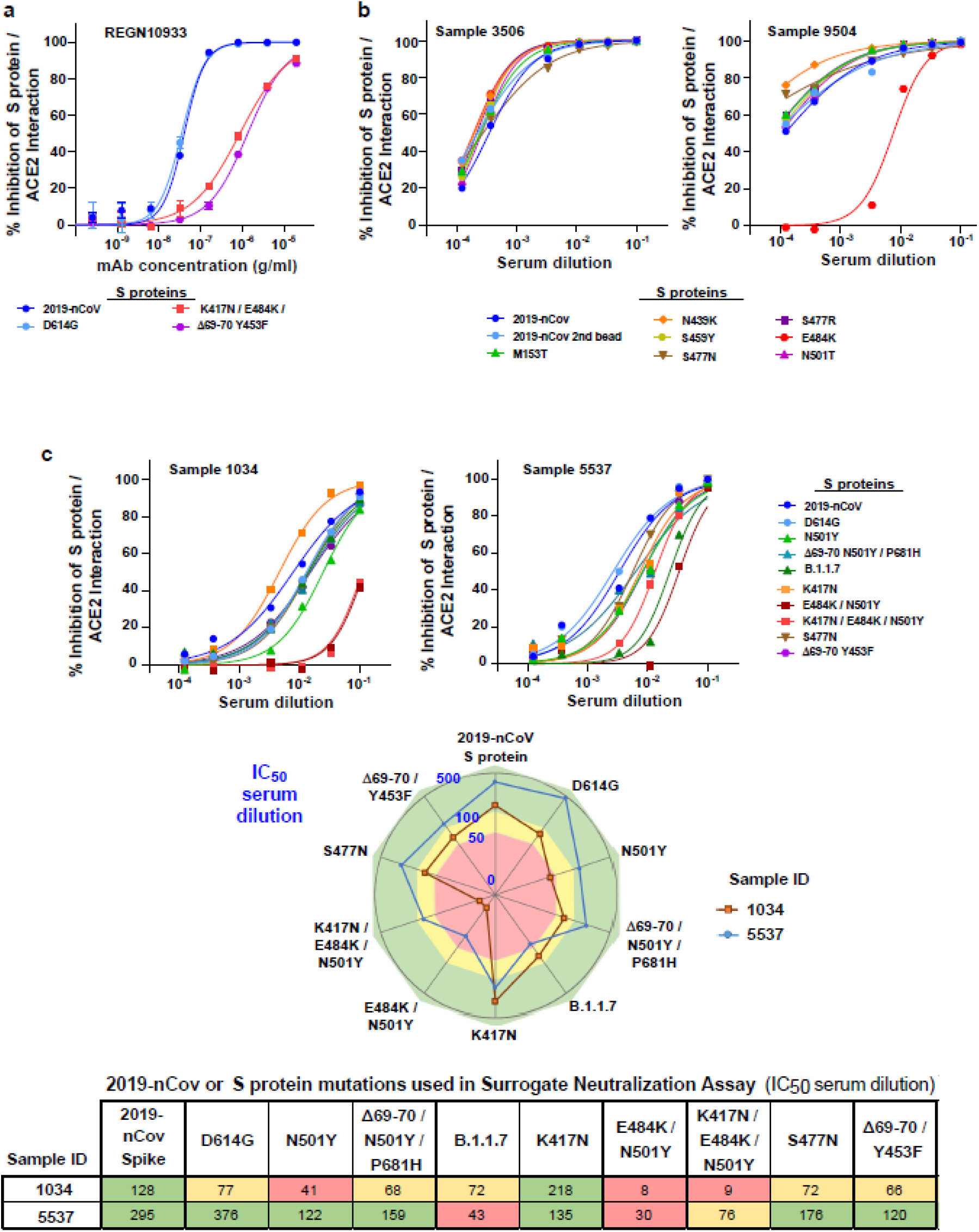
Multiplexed analysis of antibody and serum neutralizing activity against VOC S protein mutations. The surrogate neutralization assay was performed with two panels of S^3^ coupled beads consisting of the 2019-nCov S protein and S mutations produced with one or more amino acid substitutions or deletions. (**a**) Concentration response curves of the REGN10933 therapeutic antibody evaluated against S protein mutations from three viral variants in the S^3^/ACE2 assay. (**b-c**) Serum dilutions from RT-PCR positive donors 3506 and 9504 (**b**) or 1034 and 5537 (**c**) in the S^3^-ACE2 assay with two separate panels of S^3^ coupled beads. (**c**) Spider plot and heatmap showing IC_50_ serum dilutions for donors 1034 and 5537 serums samples against the indicated S protein mutations found in variants of concern including B.1.1.7 and P1. In these graphs, green background corresponds to an IC_50_ dilution >100 for strong blocking of the S^3^/ACE2 interaction, yellow background for moderate blocking with IC_50_ dilutions from 50 to 100 and red background for low to no blocking activity against the indicated S protein mutant.

**Figure 5:**
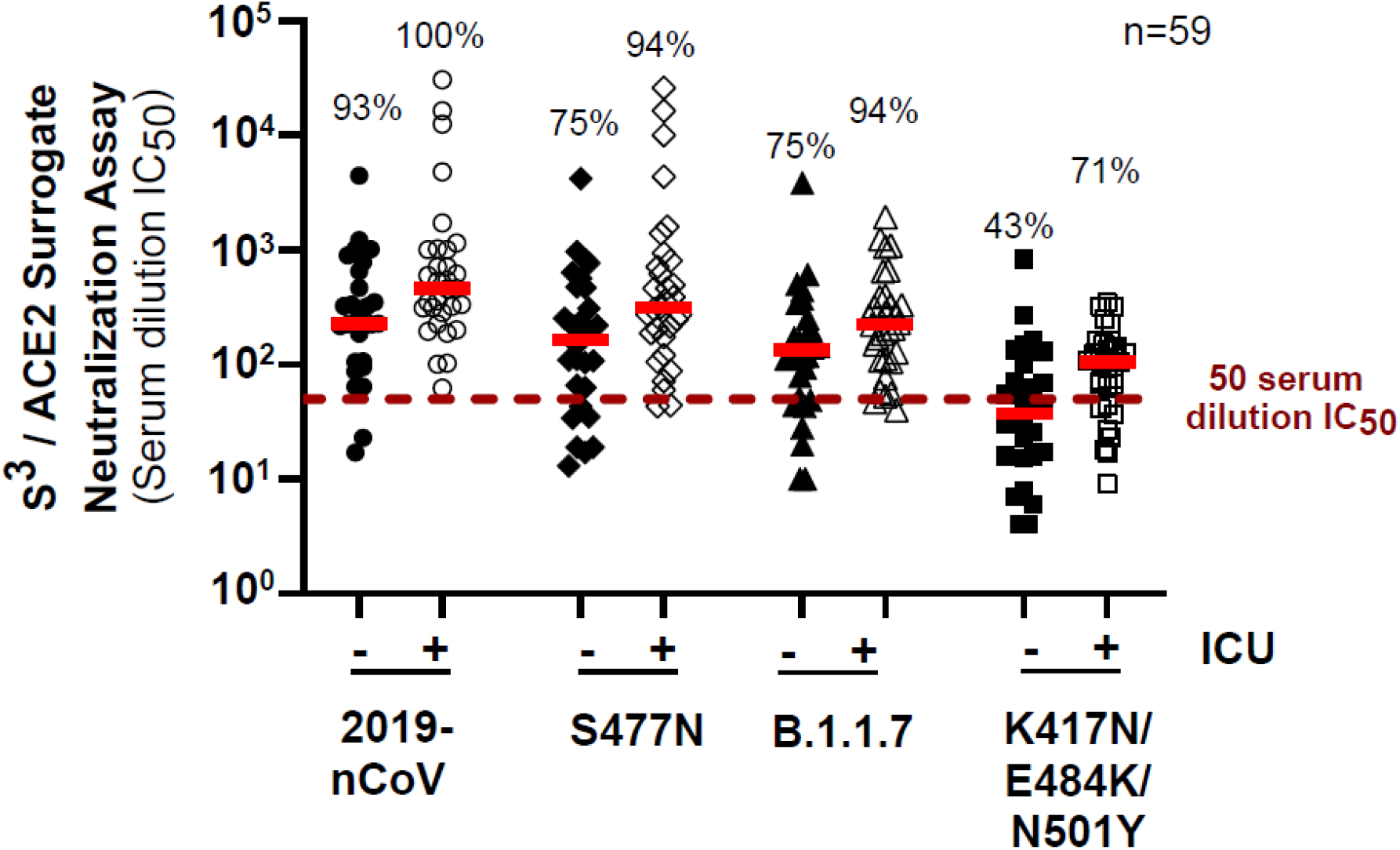
Multiplexed analysis of COVID-19 patient serum samples for neutralizing activity against variant of concern S protein mutations. Serum samples from 59 COVID-19 hospitalized patients were profiled in the multiplexed S^3^/ACE2 surrogate neutralization assay performed in parallel with the 2019-nCov S protein and three S mutants produced with one or more amino acid substitutions or deletions. S^3^/ACE2 serum dilution IC_50_ values for each of the S proteins were calculated for samples from the 28 COVID-19 hospitalized donors that remained on the wards (non-ICU, closed symbols) and for 31 COVID-19 patients that were critically ill, requiring treatment by the intensive care unit (ICU, open symbols). Dashed red line indicates the 50 serum dilution IC_50_ value cut-off in the S^3^/ACE2 assay that corresponds to neutralizing antibody levels needed for near complete (>99%) neutralization in the live virus CPE assay. Percentages of serum samples above this cut-off are shown for non-ICU and ICU donors for each of the S proteins tested. Median values are shown as a red bar for each of the data sets. Statistical analysis was performed using the non-parametric Mann-Whitney t-test and showed that ICU patients had higher levels of neutralizing antibodies compared to non-ICU patients (P=0.0127 to 0.0425) when compared for the same S protein. In comparisons between S proteins, reduced levels of neutralizing antibodies against the S protein with K417N/E484K/N501Y mutations were strongly significant (P<0.0001) relative to the 2019-nCoV S protein.

An additional panel of S^3^-coupled beads was manufactured to evaluate mutations present in the 501Y lineage circulating SARS-CoV-2 variants, including the so-called B.1.1.7 and B.1.351 or P.1 VOCs (**Figure 2b-c**). A representative donor (sample 1034) exhibited neutralizing antibody levels in the higher range of activity (>100 IC_50_ serum dilution) against the 2019-nCov S protein but only retained moderate blocking activity (50 to 100 IC_50_; yellow range in **Figure 2c**) against most of the singly or multiply mutated S protein derivatives. Importantly, this serum was almost completely inefficient against S proteins containing the E484K mutation found in the B.1.351 and P.1 VOCs (<50 IC_50_; light red range). Another serum (sample 5537) had a stronger overall neutralization activity but still displayed lower efficacy against S protein derivatives harboring the E484K mutation or against the B.1.1.7 variant (IC_50_ serum dilutions between 30 and 76).

To illustrate the high-throughput screening capability of the S^3^-ACE2 assay, neutralizing antibody titers against the 2019-CoV and three S protein variants were evaluated in serum from 59 COVID-19 patients hospitalized with (n=31) or without (n=28) need for intensive care unit (ICU) stay (**Figure 3**). The majority of patients from both subgroups, all of whom had been infected prior to November 2020, exhibited strong neutralizing activity when profiled against the 2019-nCoV S protein, with 93% of non-ICU and 100% of ICU patients having greater than 50 serum dilution IC_50_ cut-off. Based on our cross-validation studies, this cut-off value corresponds to antibody levels in undiluted serum samples that would be sufficient to achieve near complete neutralization in the live virus CPE assay (**Supplementary Figure 1**) However, they were less efficient at blocking ACE2 interaction with the S protein found in the B.1.1.7 variant or derivatives containing the S477N substitution or the K417N/E484K/N501Y triple RBD mutation encountered in the B.1.351 variant. Both groups notably exhibited a marked reduction in median IC_50_ against this latter allele (P<0.0001), with only 58% (43% of non-ICU and 71% of ICU patients) displaying antibody levels above the 50 serum IC_50_ cut-off. ICU patients additionally presented a weak but significant median increase in neutralizing antibody levels for each of the S proteins tested compared to non-ICU patients (P=0.0127 to 0.0425), consistent with a model whereby more severe and prolonged infections lead to stronger humoral immune responses.

## Discussion

Determining the level of immune protection conferred by prior SARS-CoV-2 infection, vaccination or the prophylactic administration of monoclonal antibodies is of paramount importance for informing individuals about their susceptibility to the virus, for adapting prophylactic measures to the evolving viral strains circulating in the population and, ultimately, for controlling the COVID-19 pandemic. While T-cell-based responses may contribute to this immunity,*(26)* neutralizing antibodies likely play a primary role in this process as they do for other acute viral infections, and represent the best available surrogate marker of protection. *(9, 27)* However, serum levels of SARS-CoV-2 neutralizing antibodies are so far only rarely measured owing to overwhelming technical difficulties and biosafety requirements, which preclude any sort of routine procedure or significant scale-up.

Addressing this shortcoming, we report here the development of a cell-free assay that allows for the quantitative and high-throughput evaluation of the neutralizing activity of biological samples such as serum against multiple SARS-CoV-2 variants in a single procedure taking less than three hours in a standard diagnostic laboratory. The S^3^-ACE2 assay relies on the fact that most neutralizing antibodies interfere with the binding of the viral S protein with its ACE2 receptor. Although neutralizing antibodies have been identified that recognize S protein outside of the RBD *(9)* and do not directly impact on ACE2 binding, these are rare and likely contribute minimally to antiviral immunity, as confirmed by the very high degree of correlation between our surrogate assay and its live virus cell-based reference counterpart, irrespective of the levels of neutralizing activity.*(8)* Our assay provides quantitative measures of serum neutralizing antibody levels and is further characterized by a very high degree of sensitivity and specificity (>96% and 100%, respectively, using the herein described protocol).

A significant advantage of the S^3^-ACE2 neutralization assay is its ability to evaluate multiple S variants in parallel using as little at 15 µl of serum, allowing the identification of an individual’s susceptibility to circulating and emerging SARS-CoV-2 viruses, whether after infection or vaccination. This was demonstrated with serum samples from 59 COVID-19 patients infected prior to the widespread emergence of VOCs. Importantly, only 43% of non-ICU hospitalized patients had neutralizing antibody levels greater than 50 serum dilution IC_50_ against the S protein with the K417N/E484K/N501Y mutation found in the B.1.351 variant, suggesting that many are not protected against this strain. The P.1 variant originally identified in Brazil has a similar mutation profile in the RBD and some of these non-ICU patients would be anticipated to be equally susceptible to infection by this SARS-CoV-2 variant as well. Individuals whose infection had required an ICU stay generally displayed higher levels of neutralizing antibodies against all tested S protein variants, although they too were less effective against the B1.351/P1 triple mutant.

The multiplexing of the S^3^-ACE2 neutralization assay could be increased to >40 different S alleles to tackle new VOCs. It advantageously compares with viral pseudotypes-based systems, where each S protein mutant requires production of a new batch of virions that have to be tested in separate assays, with their infectivity potentially affected by the mutations and neutralization titers influenced by the levels of ACE2 on the surface of target cells *(28, 29)*. As levels of anti-SARS-CoV-2 immunity increase in the world population due to the combined influence of ongoing infections and more widespread vaccination, selective pressures will increasingly be exerted on the virus favoring the emergence of escape mutants. The detection of these escapees should be as fast as possible for the swift adaptation of prophylactic measures including vaccines. While the infection of previously infected or vaccinated individuals will remain the strongest evidence of gaps in the collective immunity, a surveillance system based on the routine sequencing of viral isolates and the immediate testing of the susceptibility of their S protein to neutralization would constitute a far more dynamic and anticipatory approach. The S^3^-ACE2 assay, because of its ease of use, would facilitate such surveillance strategy. Furthermore, it could also be used for the high-throughput screening of candidate monoclonal antibodies and other prophylactic or therapeutic approaches aimed at blocking the interaction between SARS-CoV-2 and its cellular receptor, and it could be adapted to other viruses for which the molecular mediators of viral entry are properly characterized.

Therefore, the hereby described method stands to have an important impact in both clinical and public health settings. In this regard, immunity passports are at the forefront of current public and political discussions as possible gateways to a return to more normal social and international exchanges, as the world emerges from the COVID-19 pandemics *(30)*. They are generally thought of essentially as vaccination certificates, a concept that suffers from major shortcomings. First, such certificates would unduly exclude people that have not yet been vaccinated but endowed with strong antiviral immunity triggered by natural infection. Second, they would not be delivered to individuals that do not respond to vaccination such as primary or acquired immunodeficiency patients and/or cancer, transplant and patients with systemic inflammatory diseases receiving immunosuppressive treatments. These individuals could however be protected by passive immunization through the administration of human monoclonal antibodies, the activity of which could be quantified in their serum *(31-33)*. Third, having been vaccinated is not a guarantee of induction of optimal immunity and protection, as found every year with the flu vaccine, and the duration of vaccine-induced SARS-CoV-2 immunity is as yet unknown.

## Data Availability

The data that support the findings of this study are available from the corresponding authors

---

**Supplemental Figure 1:**
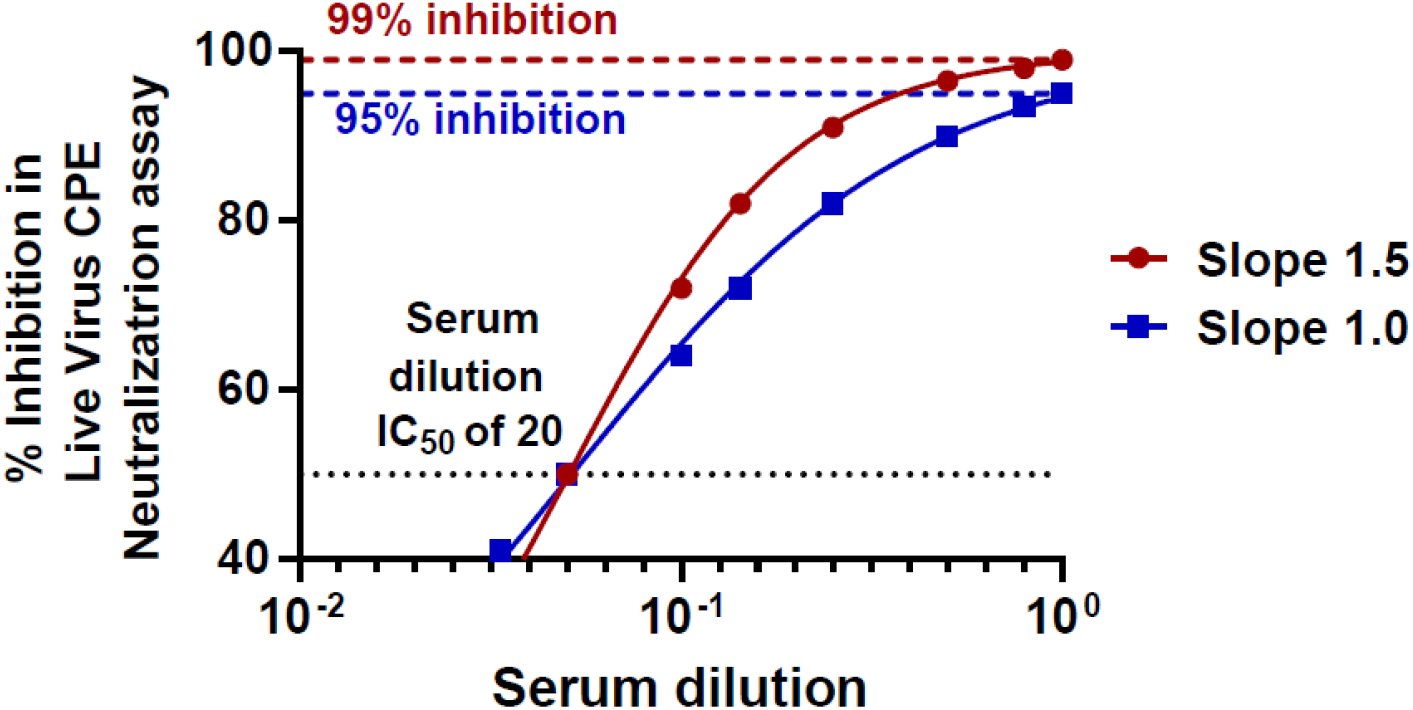
Selection of a positive cutoff values in the live virus CPE neutralization assay. Simulated data generated to illustrate that maximum viral inhibitions of 95% and 99% would be obtained with a serum dilution IC_50_ of 20 using either Hill slope values of 1.0 and 1.5, respectively.

**Supplemental Figure 2:**
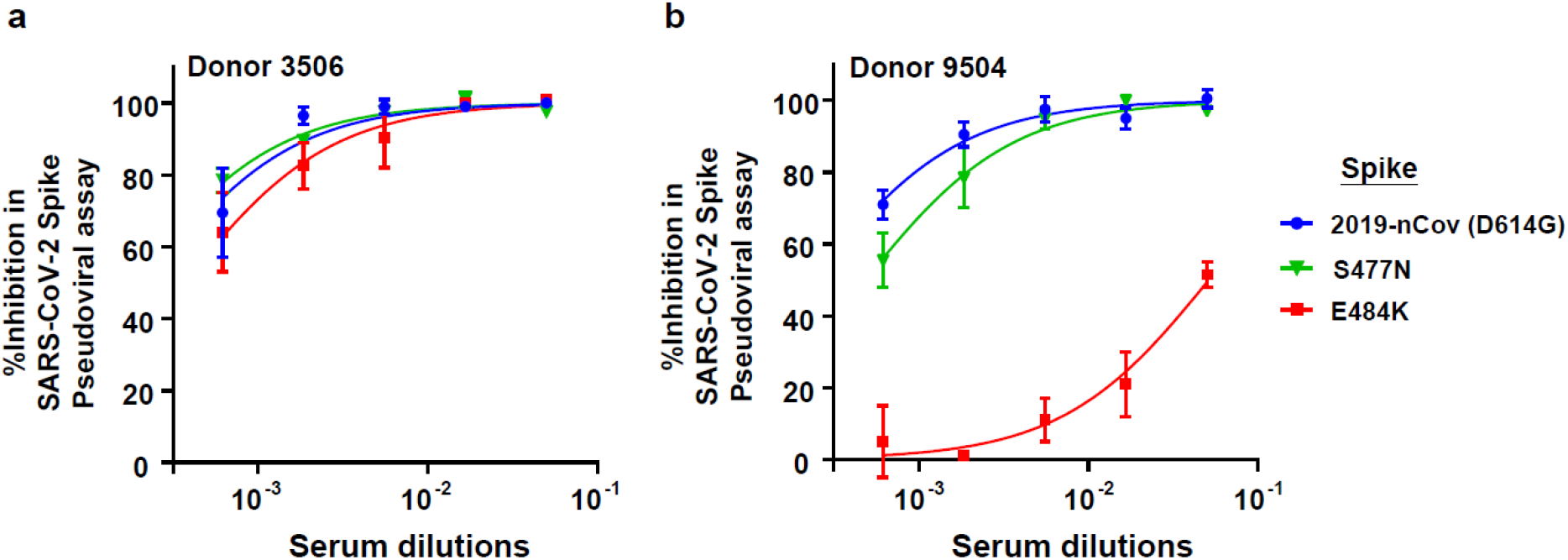
Neutralization activity of donor sera tested against pseudotyped viruses produced with different S protein mutations. Concentration response curves generated for donor 3506 (**a**) and 9504 (**b**) serum samples in the SARS-CoV-2 S protein pseudoviral assay using either the 2019-nCoV D614G, S477N/ D614G or E484K D614G S protein pseudotyped particles. Graphs show the mean of n=2 ± s.d.

**Supplemental Table 1:**
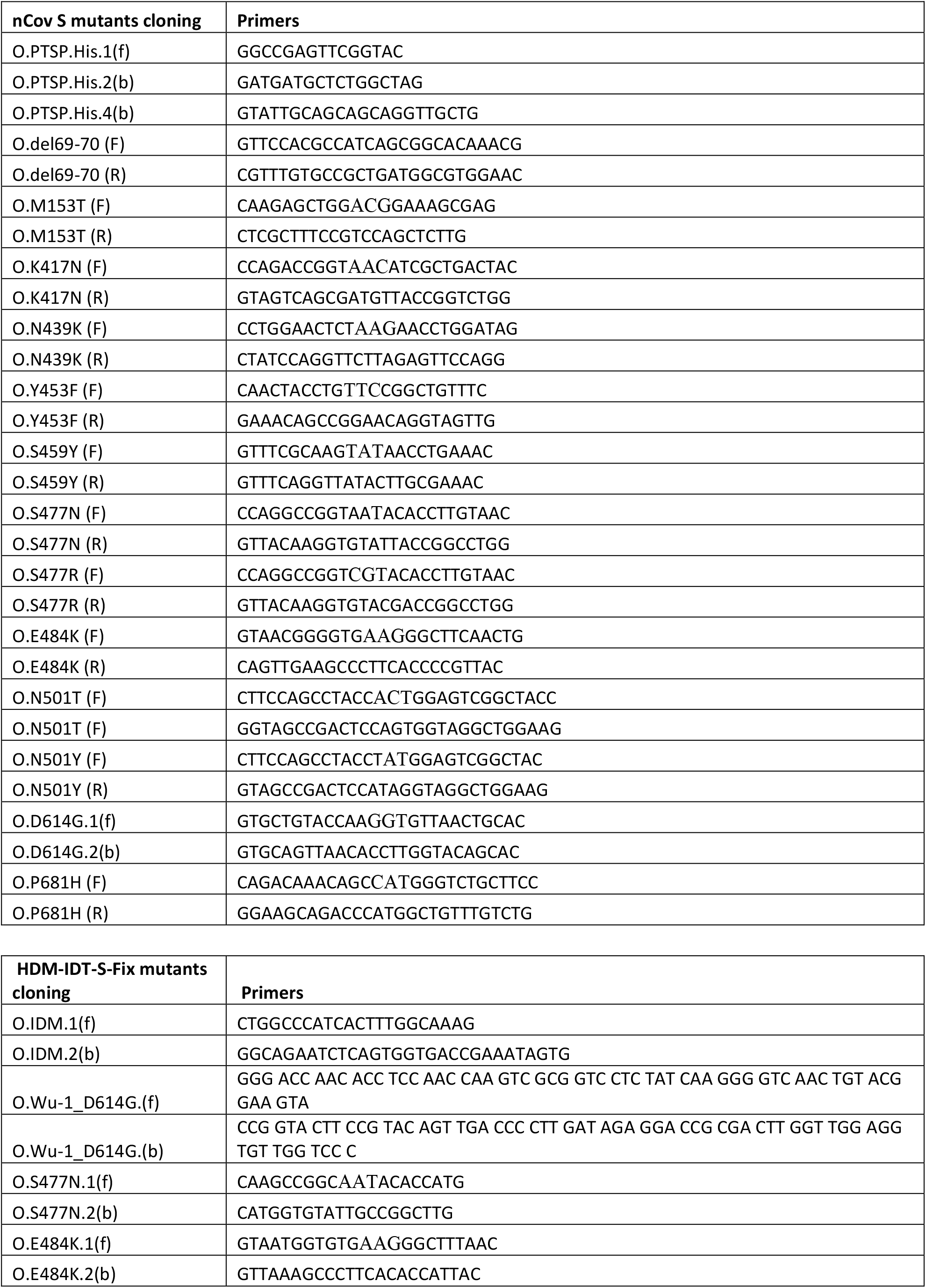
Primers used to construct Spike mutants.

## Material and Methods

### Study populations

#### Seropositive donors

Cross-validation studies were performed on serum samples identified from the seroprevalence study of the Vaud Canton in Switzerland (SerocoViD) performed by the Centre for Primary Care and Public Health, University of Lausanne (Unisanté), from the Swiss Population based seroprevalence study performed by Coronas Immunitas and from the with hospitalized donor ImmunoCov study performed by the Immunology and Allergy Service, Lausanne University Hospital. The panel of 206 SARS-CoV-2 seropositive samples consists of: 31 hospitalized COVID-19 patients and 64 RT-PCR positive non-hospitalized donors for a total of 95 RT-PCR donors and 111 seropositive donors identified through contact with RT-PCR positive donors, volunteers and asymptomatic or pausisymptomatic donors selected at random from the general population. Serum samples from 59 COVID-19 patients hospitalized with (n=31) or without (n=28) need for stay in the intensive care unit were selected from donors participating in the ImmunoCov study and Unisanté studies.

### Pre-COVID-19 pandemic donors

Negative control serum samples from 90 adult healthy donors with ages ranging for 18 to 81 years of age were collected prior to November 2019 as part of the Swiss Immune Setpoint study sponsored by Swiss Vaccine Research Institute. Study design and use of subject sera samples were approved by the Institutional Review Board of the Lausanne University Hospital and the ‘Commission d’éthique du Canton de Vaud’ (CER-VD) stated that authorization was not required.

### Production of SARS-CoV-2 S proteins

The S protein trimer was designed to mimic the native trimeric conformation of the protein in vivo and the expression vector was kindly provided by Prof. Jason McLellan, University of Texas, Austin. It encoded the prefusion ectodomain of the original 2019-Cov S protein with a C-terminal T4 foldon fusion domain to stabilize the trimer complex along with C-terminal 8x His and 2x Strep tags for affinity purification. The trimeric Spike protein was transiently expressed in suspension-adapted ExpiCHO cells (Thermo Fisher) in ProCHO5 medium (Lonza) at 5 x106 cells/mL using PEI MAX (Polysciences) for DNA delivery. At 1 h post-transfection, dimethyl sulfoxide (DMSO; AppliChem) was added to 2% (v/v). Following a 7-day incubation with agitation at 31 °C and 4.5% CO2, the cell culture medium was harvested and clarified using a 0.22 µm filter. The conditioned medium was loaded onto Streptactin (IBA) and StrepTrap HP (Cytiva) columns in tandem, washed with PBS, and eluted with 10 mM desthiobiotin in PBS. The purity of S protein trimers was determined to be > 99% pure by SDS-PAGE analysis. Generation of S protein expression vectors encoding the mutations D614G, D614G plus M153T, N439K, S477N, S477R, E484K, S459Y, N501T, N501Y, K417N, Δ60-70, P681H, Y453F or combinations thereof were generated by InFusion-mediated site directed mutagenesis using primers listed in Supplementary Table 1. The B1.1.7 variant clone was generated by gene synthesis (Twist Biosciences). S proteins for all mutants were produced and purified in an identical manner to the original 2019-Cov S protein.

### Coupling of Luminex beads with SARS-CoV-2 S protein

Luminex beads used for the serological binding assays were prepared by covalent coupling of SARS-CoV-2 proteins with MagPlex beads using the manufacture’s protocol with a Bio-Plex Amine Coupling Kit (Bio-Rad, France). Briefly, 1 ml of MagPlex-C Microspheres (Luminex) were washed with wash buffer and then resuspended in activation buffer containing a freshly prepared solution of 1-ethyl-3-(3-dimethylaminopropyl) carbodiimide (EDC) and N-hydroxysulfosuccinimide (S-NHS), (ThermoFischer, USA). Activated beads were washed in PBS followed by the addition of 50 μg of protein antigen. The coupling reaction was performed at 4 °C overnight with bead agitation using a Hula-Mixer (ThermoFischer). Beads were then washed with PBS, resuspended in blocking buffer then incubated for 30 minutes with agitation at room temperature. Following a final PBS washing step, beads were resuspended in 1.5 ml of storage buffer and kept protected from light in an opaque tube at 4 °C. Each of the SARS-CoV-2 proteins was coupled with different colored MagPlex beads so that tests could be performed with a single protein bead per well or in a multiplexed Luminex serological binding assay.

### Cell-free S protein /ACE2 surrogate neutralization assay

S protein coupled beads are diluted 1/100 in PBS with 50 µl added to each well of a Bio-Plex Pro 96-well Flat Bottom Plates (Bio-Rad). Following bead washing with PBS on a magnetic plate washer (MAG2x program), 80 µl of individual serum samples at different dilutions (e.g. 1/10, 1/30, 1/90, 1/300, 1/2700 and 1/8100) in PBS were added to the plate wells. Control wells were included on each 96-well plate that included beads alone, matching serum dilutions of a control pool of pre-COVID-19 pandemic healthy human sera (BioWest human serum AB males; VWR) and a positive control commercial anti-Spike blocking antibody (SAD-S35 from ACRO Biosciences) or recombinant produced REGN10933 neutralizing antibody discovered and marketed by Regeneron tested in a concentration response. Plates were agitated on a plate shaker for 60 minutes then the ACE2 mouse Fc fusion protein (Creative Biomart or produced by EPFL Protein Production and Structure Core Facility) was then added to each well at a final concentration of 1 µg/ml and agitated for a further 60 minutes. Beads were then washed on the magnetic plate washer and anti-mouse IgG-PE secondary antibody (OneLambda ThermoFisher) was added at a 1/100 dilution with 50µl per well. Plates were agitated for 45 minutes, washed, the beads resuspended in 80 µl of reading buffer then read directly on a Bio-Plex 200 plate reader (Bio-Rad). REGN10933 used in this study was a generous gift from Berend-Jan Bosch, Utrecht University, Netherlands. MFI for each of the beads alone wells were averaged and used as the 100% binding signal for the ACE2 receptor to the bead coupled Spike trimer. MFI from the well containing the high concentration (> 1 µg/ml) of commercial anti-Spike blocking antibody was used as the maximum inhibition signal. The percent blocking of the S protein trimer / ACE2 interaction was calculated using the formula: % Inhibition = (1-([MFI Test dilution – MFI Max inhibition] / [MFI Max binding – MFI Max inhibition]) × 100). Serum dilution response inhibition curves were generated with GraphPad Prism 8.3.0 using NonLinear four parameter curve fitting analysis of the log(agonist) vs. response. Sensitivity, specificity and correlative of the tests were calculated with Excel and GraphPad prism.

### SARS-CoV-2 live virus cell based cytopathic effect neutralization assay

All the biosafety level 3 procedures were approved by the competent Swiss authorities. The day before infection VeroE6 cells were seeded in 96-well plates at a density of 1.25×10E+4 cells per well. Heat inactivated sera from patients were diluted 1:10 in DMEM 2% FCS in a separate 96-well plate. Four-fold dilutions were then prepared in DMEM 2% FCS in a final volume of 60 µl. SARS-CoV-2 (hCoV-19/Switzerland/GE9586) viral stock (2.4×10E6/ml as titrated on VeroE6 cells) diluted 1:100 in DMEM 2% FCS was added to the diluted sera at a 1:1 volume/volume ratio. The virus-serum mixture was incubated at 37°C for 1 hour then 100 µl of the mixture was subsequently added to the VeroE6 cells in duplicates. After 48 hours of incubation at 37°C cells were washed once with PBS and fixed with 4% formaldehyde solution for 30 minutes at room temperature. Cells were washed once with PBS and plates were put at 70°C for 15 minutes for a second inactivation. Staining was performed outside the BSL3 laboratory with 50 µl of 0.1% crystal violet solution for 20 minutes at RT. Wells were washed 3 times with water and plates were dried, scanned and analyzed for the density of live violet stained cells using ImageJ software. For each 96-well plate, at least 4 wells were treated with a negative pool of sera from pre-pandemic healthy donors and 4 wells with virus only and used as negative and positive controls respectively. The percent inhibition of cytopathic effect of the virus was calculated using the formula: % Inhibition = (1-([cell density Test dilution – cell density Max inhibition] / [cell density Max binding – cell density Max inhibition]) × 100. Serum dilution response inhibition curves were generated with GraphPad Prism 8.3.0 using NonLinear four parameter curve fitting analysis of the log(agonist) vs. response. Sensitivity, specificity and correlative of the tests were calculated with Excel and GraphPad prism. Neutralization IC50 values were calculated as described above for the cell free neutralization assay using the GraphPad prism NonLinear four parameter curve fitting analysis.

### Spike-pseudotyped lentivectors production and neutralization assays

HDM-IDTSpike-fixK plasmid (a kind gift from J. Bloom) encoding for the Wuhan-Hu-1 SARS-Cov2 Spike was modified using QuickChange mutagenesis to generate the D614G mutant and D614G/S477N or D614G/E484K double mutants (primers used are listed in Table 1).Spike-pseudotyped lentivectors were generated by co-transfecting HDM-IDTSpike-fixK, pHAGE2-CMV-Luc-ZSgreen, Hgpm2, REV1b and Tat1b (a kind gift from J.D. Bloom) plasmids into 293T cells for 24 hours with the following ratio 3/9/2/2/2 (18µg/ 56.7cm2 plate) using Fugene transfection reagent (Promega). The following day, cells were transferred in EpiSerf medium, and cell supernatants were collected after 8 hours and 16 hours. Harvested supernatants were pooled, clarified by low-speed centrifugation, filtered to remove cell debris and aliquoted. Lentivector stocks were titrated and normalized for HIV antigen p24 content by ELISA (Zeptometrix).

In the pseudoviral neutralization assay, 293T cells stably expressing the ACE2 receptor were suspended in DMEM medium with 10% FCS and seeded at 1.0×10E+4 cells per well into 96-well plates. After 5 hours in cell culture at 37 °C, three-fold dilutions of serum samples were prepared and pre-incubated with the same amount of each pseudovirus in a final volume of 100 µl in DMEM + 10% FCS. Following a further 1 hour incubation at 37 °C, the pseudoviruses/serum mixture was added to the 293T ACE2 cells. After 48 hours of incubation at 37 °C, a luciferase assay was performed to monitor pseudoviral infection, using the ONE-Step™ Luciferase assay system as recommended by the manufacturer (BPS Bioscience). Viral neutralization resulted in the reduction of the relative light units detected. Neutralization IC50 values were calculated as described above for the cell free neutralization and CPE assays using the GraphPad prism NonLinear four parameters curve fitting analysis.

## Acknowledgements

We thank the Service of Immunology and Allergy at the Lausanne University Hospital for their contributions in analysis of subject serum samples for levels of anti-S protein IgG and IgA antibodies, Laurence Durrer and Soraya Quinche from PTPSP-EPFL for mammalian cell culture, Michaël François and Kelvin Lau from PTPSP-EPFL for helping in purification of the S protein trimer proteins and ACE2-Fc protein. We would like to thank the Trono laboratory, Caroline Tapparel for the help to start the SARS-Cov2 related work and I. Eckerle for providing a SARS-Cov2 isolate. We also thank Valerie D’Acremont from Centre for Primary Care and Public Health, University of Lausanne, in Lausanne, Switzerland and Milo Puhan and Jan Fehr from the University Hospital of Zurich for providing serum samples from the Unisanté and Swiss Population based seroprevalence study performed by Coronas Immunitas, respectively. Funding for this project was provided through the Lausanne University Hospital, through the Swiss Vaccine Research Institute and through the Coronavirus Accelerated R&D in Europe (CARE) IMI project to G.P., and through the EPFL COVID fund to D.T.

## Author contributions

P.T. established and performed the live SARS-CoV-2 virus cytopathic effect neutralization assay, designed the S protein mutations and cloning, analyzed the results and contributed to the editing of the manuscript. C.F. conceived of and designed the S protein /ACE2 surrogate neutralization assays, organized the testing of sera, analyzed the data, wrote the initial draft and contributed to the editing of the manuscript. C.P. optimized the conditions for the S protein /ACE2 surrogate neutralization assays, solidified the testing protocol into a robust diagnostics assay and tested all the serum samples with this assay. A.F. prepared all batches protein coupled Luminex beads. J.C. performed all pseudoviral assays; C.R. performed site directed mutagenesis of the S protein constructs; F.P. and the Protein Production and Structure Core Facility at the EPFL produced and purified the trimer S proteins. V.C. provided initial stocks of titrated SARS-CoV-2 virus. D.T. and G.P. conceived the study design for testing of sera from the different subject groups, analyzed the results and wrote the manuscript.

